# Developing a Neuropsychiatry Curriculum for Clinical Psychologists and Neuropsychologists: An e-Delphi Study

**DOI:** 10.64898/2026.05.14.26353190

**Authors:** Keishema Kerr, Tamara Anderson, Graham Blackman, Amy Copping, Niels Detert, Alexandra Garfield, Peter Gilli, Laura Goldstein, Huw Green, Simon Harrison, Leigh Leppard, Norman Poole, Torie Robinson, Alexandra Rose, Biba Stanton, Mary Summers, Victoria Teggart, Mike Wang, Vaughan Bell

**Affiliations:** Clinical, Educational and Health Psychology, University College London, UK; Lewisham Acute and Crisis Psychology, South London and Maudsley NHS Foundation Trust, UK; Department of Psychiatry, University of Oxford, Warneford Hospital, Oxford, UK; Oxford Health NHS Foundation Trust, Oxford, UK; West Kent and Medway Neuropsychiatry Team, Kent and Medway Mental Health NHS Trust, UK; Russell Cairns Unit, Adult Neuropsychology, Oxford University Hospitals NHS Foundation Trust, UK; Kent Clinical Neuropsychology Service, Kent and Medway Mental Health NHS Trust, UK; FND Portal, USA; Dept of Psychology, Institute of Psychiatry, Psychology and Neuroscience, King’s College London, UK; Department of Neuropsychology, Addenbrooke’s Hospital, Cambridge, UK; Department of Neuropsychiatry, South London and Maudsley NHS Foundation Trust, UK; Lishman Unit, South London and Maudsley NHS Foundation Trust, UK; Epilepsy Sparks Insights, UK; Royal Hospital for Neuro-disability, London, UK; School of Health and Wellbeing, College of Medical, Veterinary and Life Sciences, University of Glasgow, Glasgow, UK; Department of Neurology, King’s College Hospital NHS Foundation Trust, UK; Greater Manchester Mental Health NHS Foundation Trust, UK; School of Psychology and Vision Sciences, University of Leicester, UK

**Author notes:** **Corresponding author:** Vaughan Bell.

**Keywords:** neuropsychiatry, training, education, professional development, Delphi

## Abstract

**Objective:** Neuropsychiatric presentations are common across neurological and mental health services but they are often inadequately covered by core clinical psychology and clinical neuropsychology training. Consequently, we aimed to identify components for a neuropsychiatry curriculum for clinical psychologists using a Delphi process.

**Method:** We completed a three-round e-Delphi study with 19 experts (clinical psychologists, neuropsychologists, psychiatrists, neurologists, individuals with lived experience of neuropsychiatric disorders). Round 1 collected ratings on 80 syllabus items derived from textbook reviews, conference topics, and a scoping review of neuropsychiatry syllabuses. Items failing to reach consensus were refined, and new topics added via free-text suggestions. Rounds 2 and 3 repeated rating and thematic analysis, culminating in a consensus meeting where items were classified as core or supplementary. Consensus thresholds were set at mean ≥ 2.0, mean distance from the mean ≤ 0.2, and ≥ 75% agreement for final decisions.

**Results:** The process yielded 40 core and 38 supplementary syllabus items. Core topics include autoimmune and neuroinflammatory disorders, delirium, functional neurological disorders, neuropsychiatric sequelae of epilepsy, stroke, traumatic brain injury, dementia, and multidisciplinary working, among others. Supplementary items covered background knowledge of less frequent but still prevalent disorders as well as competencies in interpreting clinical data alongside conceptual and historical issues. The final component list reflects both clinical competencies and emerging areas of practice, emphasising assessment, formulation, psychological interventions, cultural considerations, and medicolegal aspects.

**Conclusions:** The e-Delphi-derived curriculum provides a framework for neuropsychiatric competencies for postgraduate psychology training with modification needed for application in diverse healthcare settings.

## Introduction

Neuropsychiatry is a clinical specialty at the intersection of neurology and psychiatry that focuses on the psychiatric and behavioural manifestations of neurological disorders and ‘cross-over’ disorders such as tic disorders and functional neurological disorders with both neurological and psychiatric characteristics (Arciniegas & Kaufer, 2006). Specialist neuropsychiatric services have been developed to provide consultation and management for complex cases (Trapp et al., 2022; Vaishnavi et al., 2009) but neuropsychiatric competencies are relevant beyond specialist services. Indeed, neuropsychiatric presentations are common across neurological and mental health services more broadly (Carson et al., 2000; Hesdorffer, 2016), suggesting that a foundational understanding of neuropsychiatric principles is essential for clinicians across disciplines.

The need for clinical psychology expertise in neuropsychiatry is supported by both patient demand and healthcare policy. Hansen et al. (2005) found that one-third of neurological patients requested additional psychological support. Psychological support or intervention has been identified as a key unmet care need in epilepsy (Boele et al., 2025; Gandy et al., 2021), dementia (Koh et al., 2025), stroke (Zawawi et al., 2020) and traumatic brain injury (Andelic et al., 2021). The Neurological Alliance (2019) recommended a dual-pronged approach to treating people with neurological conditions, integrating both medical and psychological aspects.

However, the extent to which neuropsychiatric competencies are included in either general clinical psychology or specialist clinical neuropsychology training varies considerably (Heffelfinger et al., 2022; Kosmidis et al., 2022; Krause et al., 2025). The place of neuropsychiatry in psychology training curriculums appears to range from absent (Sheer & Lubin, 1980), implied but not explicitly included (Hessen et al., 2018; Wong et al., 2024), narrowly defined as neurodevelopmental disorders (Hokkanen et al., 2022), or where expertise in ‘neuropsychiatric disorders’ is included as an explicitly required competency but without further elaboration (BPS Division of Neuropsychology, 2012; Nelson et al., 2015).

Consequently, we set out to define a neuropsychiatry curriculum for clinical psychologists and neuropsychologists using the e-Delphi method, an iterative, consensus-building approach that systematically gathers expert opinion through multiple survey rounds (Hsu & Sandford, 2007). The Delphi method is considered appropriate when there is limited established guidance on a topic, and expert consensus is required to inform best practice or curriculum development (Keeney et al., 2011). It is particularly suited to complex, multidisciplinary fields such as neuropsychiatry, where perspectives from diverse professional groups are valuable for generating a comprehensive and balanced framework and has been widely used for curriculum development in clinical practice (e.g. Connery et al., 2022; Viljoen et al., 2020). Our Delphi approach included clinical psychologists and neuropsychologists working in neuropsychiatry, alongside colleagues from psychiatry and neurology, and individuals with lived experience of neuropsychiatric services to define a core neuropsychiatry curriculum for psychologists.

## Methods

This study was approved by the University College London ethics committee (Ref: CEHP/2023/594).

### Delphi procedure

A modified three-round e-Delphi methodology was employed to achieve consensus among an expert panel (Woodcock et al., 2020). Rounds one and two involved anonymous collection of expert ratings and qualitative feedback, followed by item selection using analysis of both quantitative and qualitative responses, and the redistribution of summary results to participants in subsequent rounds. This iterative feedback process enables panel members to reconsider their responses in light of group trends while maintaining anonymity, thus reducing the influence of dominant individuals and promoting genuine consensus (Hsu & Sandford, 2007). The e-Delphi method is widely recognised as best practice for developing expert opinions on novel topics and is the preferred method for curricular recommendations (Sharma et al., 2019). Three-round processes are preferred in Delphi studies, as evidence suggests it minimises research fatigue and produces stable results, with the most significant adjustments typically occurring between rounds one and two (Boulkedid et al., 2011).

The process of generating the initial items for the modified e-Delphi survey was informed by a review of neuropsychiatry and neuropsychology textbooks, topic lists from recent conferences, and a scoping review of neuropsychiatry syllabuses (Kerr et al., 2025).

We used two online survey rounds and, a final consensus meeting conducted as a live video conference. Participants were instructed “Please assume that you are selecting a syllabus that would fit into a year’s worth of part-time post-qualification teaching to be conducted alongside clinical work to ensure adequate competencies in neuropsychiatry for qualified clinical psychologists. Assume candidates would already be competent in psychological interventions for common mental health problems and assessment and interventions for cognitive problems after common neurological disorders” to ensure they were selecting for additional competencies not already covered by core training.

For the two online survey rounds, panel members rated syllabus items using a three-point Likert scale: 1. “Not necessary to include”, 2. “Supplementary syllabus (to be understood in outline)” and 3. “Core syllabus (to be understood in-depth)”. The three-point Likert scale was selected for its ability to produce more accurate consensus ratings and its strong test-retest reliability (Lange et al., 2020). Mean, median, and Mean Distance from the Mean (MDM) were calculated for each item. Consensus for inclusion was defined as mean ≥2.0 and MDM ≤0.2 (Keeney et al., 2011; Murry & Hammons, 1995). Items with >50% rejection rate were excluded (Fink et al., 1984). Participants could suggest additional topics via free-text responses which were included in the next round. For the final meeting, conducted as an online video conference, items achieving preliminary consensus (median ≥2.0, MDM ≤0.2) were presented for final categorisation. Items lacking clear consensus were prioritized for discussion. Final decisions required ≥75% panel agreement (Jünger et al., 2017).

### Selecting the Panel

Expert panel members are defined as those with in-depth knowledge and / or experience in the field (de Villiers et al., 2005). Panel members were recruited through professional networks and invited based on their experience working in neuropsychiatry or neuropsychiatric services. A deliberately diverse panel was invited, covering clinical psychology, neuropsychology, psychiatry, neurology, and individuals with lived experience as patients in neuropsychiatric services. The target was to recruit 20 panel members, allowing for potential attrition, with 10-15 participants considered sufficient for e-Delphi studies (Hong et al., 2019), and 19 finally recruited for the study.

## Results

### Panel members

Panel members who participated in round one (N=19) were predominantly female (N=11) and had an average of 14.5 (SD = 10.2) years of experience in neuropsychiatry. Participants were clinical psychologists (N=6), clinical neuropsychologists (N=6), neuropsychiatrists (N=3), individuals with lived experience of neuropsychiatry services and disorders (N=2), a neurologist (N=1), and an academic clinical neuropsychologist (N=1). Participants reported working in the following services: neuropsychiatry services, including both inpatient and outpatient settings (N=6); neurorehabilitation services, including those supporting individuals with acquired brain injury, prolonged disorders of consciousness, and functional neurological disorder (N=3); neuropsychology services in acute hospital and rehabilitation contexts (N=3); acute adult mental health services (N=1); general neurology services, including FND (N=1); addictions and homelessness services (N=2); solely in education or university department (N=1); and roles based on patient advocacy or lived experience (N=2).

### Delphi procedure

The flowchart illustrating the Delphi procedure is displayed in Figure 1.

**Figure 1.**
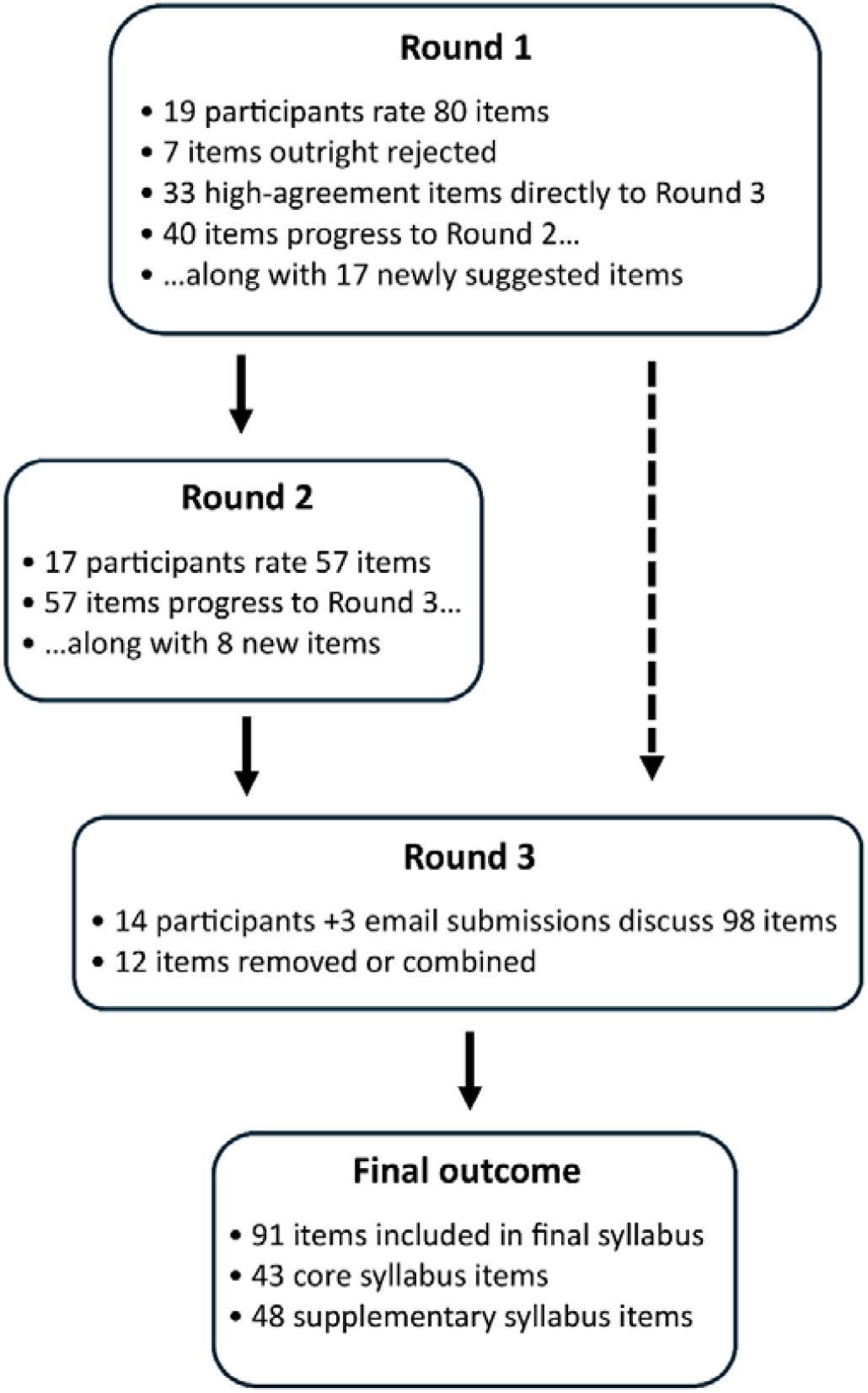
Flowchart for Delphi procedure

#### Round one results

All median and mean distance measures (MDMs) for the 80 syllabus items rated in round one are displayed in Table S1 of the supplementary material. Seven items were excluded prior to round two due to meeting the rejection criterion of >50% of respondents rating the item as “not necessary for inclusion” (full ratings in Table S2 of the supplementary material). These were synaesthesia, disorders of smell, interpreting EEG results, interpreting genetic tests, body integrity identity disorder, interpreting blood test results, and ‘art therapy techniques and applications’. In total, 33 topics (Table S4, supplementary material) reached the high agreement consensus criteria (median rating >2 and an MDM ≤0.2) and advanced directly to round three. Panel members proposed 17 new topics for consideration via the free-text box (full items in Table S3, supplementary material). These included additions reflecting a broadening of the syllabus to encompass practical, legal, and interdisciplinary aspects of neuropsychiatric practice, alongside emerging and underrepresented clinical topics such as COVID-19 related neuropsychiatry, subtle symptom presentations, and the neuropsychological dimensions of both neurological and psychiatric disorders. In addition, one panel member also proposed combining the mental capacity with the frontal lobe paradox item.

#### Round two results

In round two, panel members rated 57 items, which included 40 items that did not meet consensus in round one and 17 items suggested during the round one free-text section. The full ratings are presented in Table S5 of the supplementary material. No items met the outright rejection criteria in round two. Five additional items reached consensus during this round, including psychotherapy for neuropsychiatric patients, qualitative aspects of performance on neuropsychological tests in neuropsychiatric syndromes, driving in the context of seizures and non-epileptic attack disorder, subtle/attenuated presentations of neuropsychiatric phenomena, and mass psychogenic illness. The remaining 52 items did not achieve consensus, though some, such as confabulation and the neuropsychiatry of COVID-19, came close to meeting the preset threshold. These items formed the basis for discussion during the consensus meeting in round three. Eight additional items were suggested in the free-text box, including social aspects of neuropsychiatric disorders, the impact of neuropsychiatric disorders on families, “health prevention” (sic), cultural variances in clinical presentation, patient awareness of seemingly unrelated symptoms, psychedelic treatment, hypnosis, and neurodivergence and how this impacts clinical presentation.

#### Round three results

The goal of the consensus meeting was to resolve outstanding items, refine the list, and finalise decisions regarding the structure of the syllabus. Prior to the meeting, respondents were provided with a proposed syllabus that categorised items as core or supplementary based on the results to date. They were invited to submit feedback, and three respondents who could not attend the meeting sent their comments via email. In addition to these comments, items with lower agreement were prioritised for discussion during the meeting. Fourteen respondents attended the consensus meeting, which was moderated by a researcher. Final decisions were made by a vote, with a predetermined consensus threshold of ≥75% agreement. Following the consensus discussions, five items were reclassified as core content, including autism, attention-deficit hyperactivity disorder, motor neuron disease, autoimmune conditions, and cerebellar syndromes (previously listed as cerebellar cognitive affective syndrome). Six topics were removed from the syllabus to streamline the curriculum. These were physiotherapy (considered outside the scope of neuropsychiatric training), frontal lobe paradox, epigenetics, impulse control disorders (deemed to be covered under other topics), post-concussion syndrome (covered under traumatic brain injury), and COVID-19 (considered under broader medical neuropsychiatric conditions). Throughout the meeting, several topics were rephrased to improve clarity and inclusivity. The topic “Multidisciplinary working for neurorehabilitation goals” was revised to the broader term “Multidisciplinary (MDT) working”, and the item “Psychotherapy for neuropsychiatric patients” was expanded to “Psychological interventions and psychotherapy for neuropsychiatric patients” to encompass a broader range of therapeutic approaches. Additionally, all content related to medications was consolidated into a single item: “Effects, adverse effects, and neuropsychiatric syndromes associated with medication used in neuropsychiatry”. Similarly, topics related to mental capacity, safeguarding, and mental health law were combined into a single item. The final core and supplementary items, as agreed upon by the panel, are presented in Tables 1 and 2.

**Table 1.**
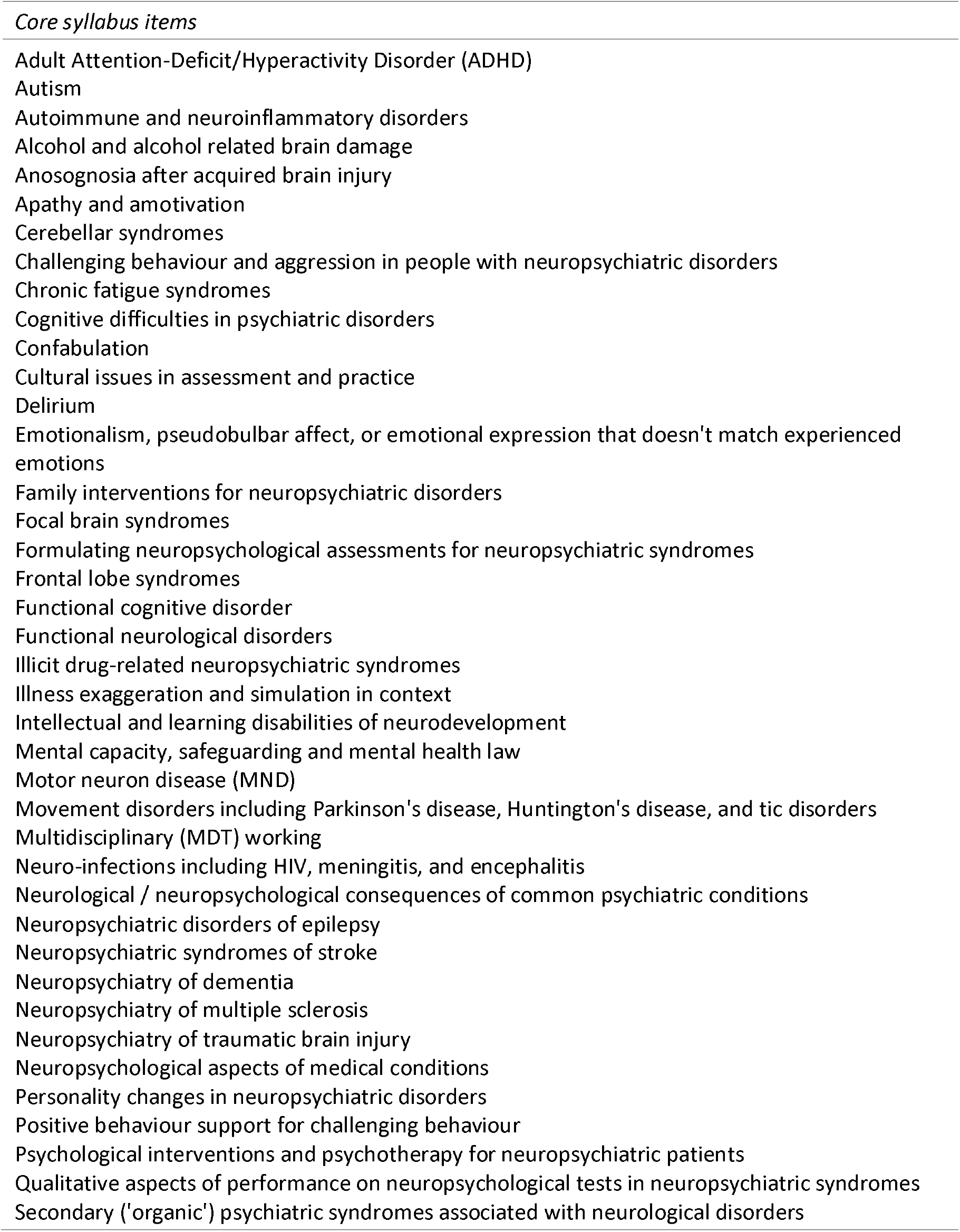
Core syllabus items finalised in e-Delphi round three consensus meeting.

**Table 2.**
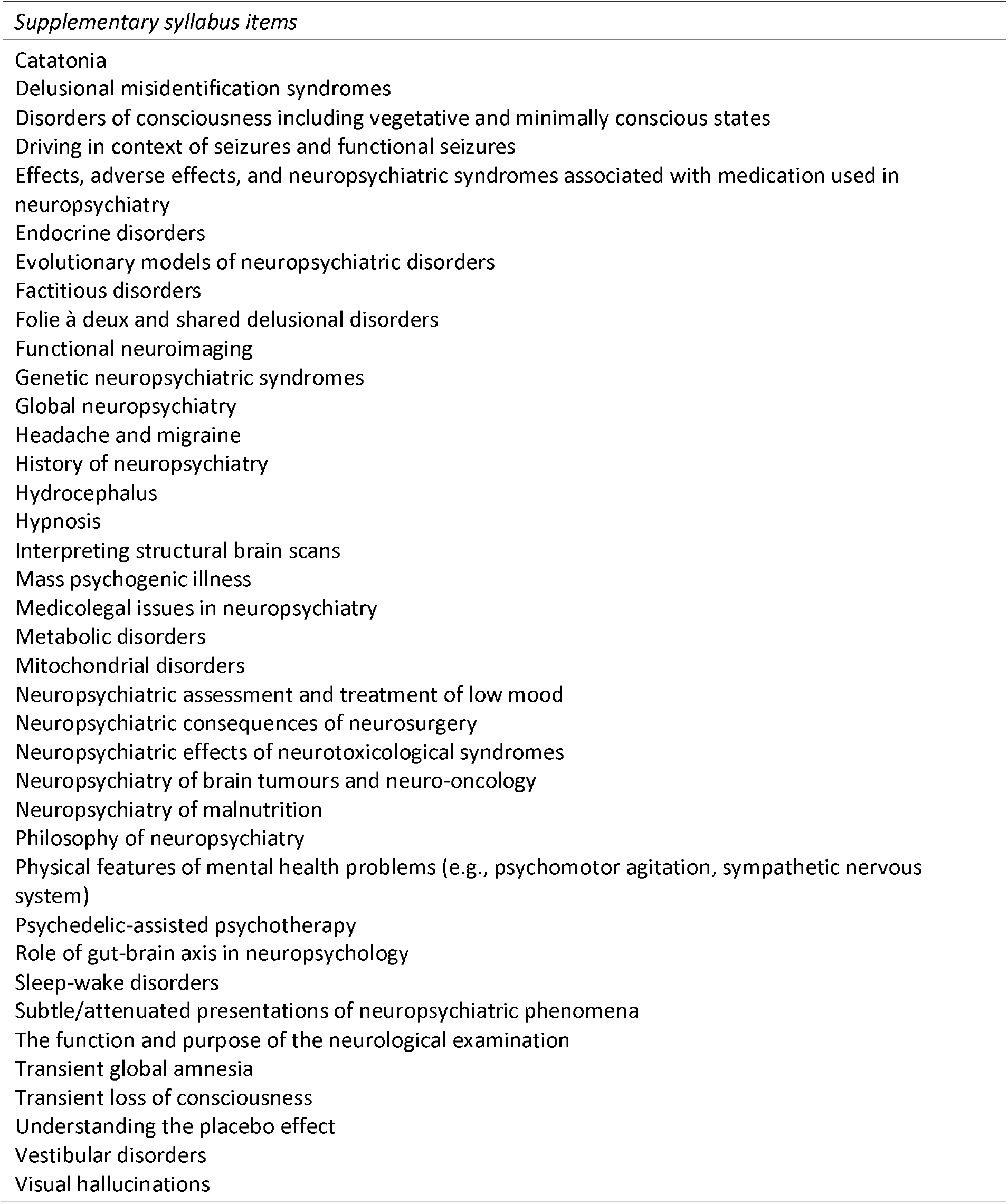
Supplementary syllabus items finalised in e-Delphi round three consensus meeting.

## Discussion

The present study outlined a neuropsychiatry syllabus for clinical psychologists and clinical neuropsychologists using an e-Delphi process to develop consensus across psychologists, medical professionals and individuals with lived experience of neuropsychiatric disorders. The syllabus consists of 40 core and 38 supplementary items intended to inform foundational competencies for working with individuals with neuropsychiatric difficulties and disorders.

In terms of core disorders, the suggested syllabus strongly overlaps with elements from a recent scoping review of international neuropsychiatry and behavioural neurology syllabuses (Kerr et al., 2025), suggesting strong validity in terms of domain coverage. However, it also includes some key psychological competencies. Notably, family interventions for neuropsychiatric disorders, formulating neuropsychological assessments for neuropsychiatric syndromes, and positive behaviour support for challenging behaviour, were not considered to be adequately adapted to the specific needs of neuropsychiatric disorders in core training and so were prioritised for inclusion in the syllabus. Similarly, while cultural issues in assessment and practice are widely covered in core training, field-specific issues – such as diversity in terms of presentation and causes of neuropsychiatric syndromes – were considered important enough to include as a dedicated item in a neuropsychiatry-specific syllabus.

However, the syllabus items, in their majority, do not describe entirely new interventions or assessment approaches that are not covered by core training, suggesting that the ‘tools’ of clinical psychologists and clinical neuropsychologists remain largely relevant to work in neuropsychiatry services. Indeed, the majority of the syllabus items related to specific neuropsychiatric presentations that were considered not to be adequately covered in core training (e.g. “Neuropsychiatric disorders of epilepsy”; Tolchin et al., 2020) or not covered in sufficient depth to address their significance and diversity in neuropsychiatric presentations (e.g. “Autism”; Khachadourian et al., 2023). Some represented a wider understanding of interventions and assessments typically carried out by non-psychologists (e.g. “Interpreting structural brain scans”, “The function and purpose of the neurological examination”), reflecting the close multidisciplinary team work common in neuropsychiatry (Taslim et al., 2024).

We also note some newer or less common topics that appeared in the supplementary syllabus list. “Psychedelic-assisted psychotherapy” is not a common treatment in neuropsychiatry services although knowledge of its practice may become increasingly important as psychedelic therapy becomes more widely available, even if largely through private clinics (Reiff et al., 2020). Similarly, “Hypnosis” is not frequently offered as an intervention although there is renewed interest in its use as a treatment for functional neurological disorder (Connors et al., 2024) and it is considered to share some mechanisms for dissociation more broadly (Bell et al., 2011).

However, we also highlight some limitations to this study. Professionals were largely drawn from the United Kingdom and most had trained, or were working within the UK National Health Service (NHS), meaning the extent to which the syllabus generalises to other countries or healthcare settings is unclear. Similarly, although there was a strong overlap between core syllabus items identified in a review of international neuropsychiatry syllabuses, neither approach accounts for regional variation where expertise in specific problems are essential to neuropsychiatric practice in some locations but less relevant in others (e.g. Ramírez Bermúdez et al., 2026). Consequently, this e-Delphi study should be considered as generating an informative rather than a definitive syllabus for psychologists working in neuropsychiatry given the need for adaptation to specific healthcare contexts.

We also note possible limitation in the process of expert selection which was largely conducted through the professional networks of the senior authors. Although this is a common selection approach, the reliance on existing networks may have introduced selection bias, potentially limiting the diversity of perspectives and under-representing clinicians or stakeholders outside those circles. Future work could aim for wider recruitment strategies, such as systematic outreach through professional societies, open calls for participation, and snowball sampling, to maximise the range of panel members.

In summary, we report an e-Delphi study aimed at outlining components for a neuropsychiatry curriculum for clinical psychologists and neuropsychologists. Given the expanding role of psychologists across health services, an increasingly ageing population, and the need for psychologists to work across traditional service boundaries, we hope this will provide a framework for training in this area.

## Data Availability

All data produced in the present work are contained in the manuscript

## References

Andelic, N., Røe, C., Tenovuo, O., Azouvi, P., Dawes, H., Majdan, M., Ranta, J., Howe, E. I., Wiegers, E. J. A., Tverdal, C., Borgen, I., Forslund, M. V., Kleffelgaard, I., Dahl, H. M., Jacob, L., Cogné, M., Lu, J., von Steinbuechel, N., & Zeldovich, M. (2021). Unmet Rehabilitation Needs after Traumatic Brain Injury across Europe: Results from the CENTER-TBI Study. Journal of Clinical Medicine, 10(5), 1035. 10.3390/jcm10051035

Arciniegas, D. B., & Kaufer, D. I. (2006). Core Curriculum for Training in Behavioral Neurology & Neuropsychiatry. The Journal of Neuropsychiatry and Clinical Neurosciences, 18(1), 6–13. 10.1176/jnp.18.1.6

Bell, V., Oakley, D. A., Halligan, P. W., & Deeley, Q. (2011). Dissociation in hysteria and hypnosis: Evidence from cognitive neuroscience. Journal of Neurology, Neurosurgery & Psychiatry, 82(3), 332–339. 10.1136/jnnp.2009.199158

Boele, F., Jensen, C., Johnson, G. M., Lammers-Spijker, A., Altinbas, A., Berg, L. van den, Broekman-Labinac, K., Fronczek, R., Schuur, M., Vonk, G., Zijlmans, M., Visser, G., & Reijneveld, J. (2025). Health-related quality of life and unmet needs of people with epilepsy and their family caregivers: A systematic scoping review. Epilepsy & Behavior, 171. 10.1016/j.yebeh.2025.110601

Boulkedid, R., Abdoul, H., Loustau, M., Sibony, O., & Alberti, C. (2011). Using and Reporting the Delphi Method for Selecting Healthcare Quality Indicators: A Systematic Review. PLOS ONE, 6(6), e20476. 10.1371/journal.pone.0020476

BPS Division of Neuropsychology. (2012). Competency framework for the UK Clinical Neuropsychology profession. British Psychological Society.

Carson, A. J., Ringbauer, B., MacKenzie, L., Warlow, C., & Sharpe, M. (2000). Neurological disease, emotional disorder, and disability: They are related: a study of 300 consecutive new referrals to a neurology outpatient department. Journal of Neurology, Neurosurgery & Psychiatry, 68(2), 202–206. 10.1136/jnnp.68.2.202

Connery, A., Yaruss, J. S., Lomheim, H., Loucks, T. M., Galvin, R., & McCurtin, A. (2022). Obtaining consensus on core components of stuttering intervention for adults: An e-Delphi Survey with key stakeholders. International Journal of Language & Communication Disorders, 57(1), 112–127. 10.1111/1460-6984.12680

Connors, M. H., Quinto, L., Deeley, Q., Halligan, P. W., Oakley, D. A., & Kanaan, R. A. (2024). Hypnosis and suggestion as interventions for functional neurological disorder: A systematic review. General Hospital Psychiatry, 86, 92–102. 10.1016/j.genhosppsych.2023.12.006

de Villiers, M. R., de Villiers, P. J. T., & Kent, A. P. (2005). The Delphi technique in health sciences education research. Medical Teacher, 27(7), 639–643. 10.1080/13611260500069947

Fink, A., Kosecoff, J., Chassin, M., & Brook, R. H. (1984). Consensus methods: Characteristics and guidelines for use. American Journal of Public Health, 74(9), 979–983.

Gandy, M., Modi, A. C., Wagner, J. L., LaFrance Jr, W. C., Reuber, M., Tang, V., Valente, K. D., Goldstein, L. H., Donald, K. A., Rayner, G., & Michaelis, R. (2021). Managing depression and anxiety in people with epilepsy: A survey of epilepsy health professionals by the ILAE Psychology Task Force. Epilepsia Open, 6(1), 127–139. 10.1002/epi4.12455

Hansen, M. S., Fink, P., Søndergaard, L., & Frydenberg, M. (2005). Mental illness and health care use: A study among new neurological patients. General Hospital Psychiatry, 27(2), 119–124. 10.1016/j.genhosppsych.2004.10.005

Heffelfinger, A. K., Janecek, J. K., Johnson, A., Miller, L. E., Nelson, A., & Pulsipher, D. T. (2022). Competency-based assessment in clinical neuropsychology at the post-doctoral level: Stages, milestones, and benchmarks as proposed by an APPCN work group. The Clinical Neuropsychologist, 36(6), 1209–1225. 10.1080/13854046.2020.1829070

Hesdorffer, D. C. (2016). Comorbidity between neurological illness and psychiatric disorders. CNS Spectrums, 21(3), 230–238. 10.1017/S1092852915000929

Hessen, E., Hokkanen, L., Ponsford, J., Zandvoort, M. van, Watts, A., Evans, J., & Haaland, K. Y. (2018). Core competencies in clinical neuropsychology training across the world. The Clinical Neuropsychologist. https://www.tandfonline.com/doi/abs/10.1080/13854046.2017.1413210

Hokkanen, L., Jokinen, H., Rantanen, K., Nybo, T., & Poutiainen, E. (2022). Status of Clinical Neuropsychology Training in Finland. Frontiers in Psychology, 13. 10.3389/fpsyg.2022.860635

Hong, Q. N., Pluye, P., Fàbregues, S., Bartlett, G., Boardman, F., Cargo, M., Dagenais, P., Gagnon, M.-P., Griffiths, F., Nicolau, B., O’Cathain, A., Rousseau, M.-C., & Vedel, I. (2019). Improving the content validity of the mixed methods appraisal tool: A modified e-Delphi study. Journal of Clinical Epidemiology, 111, 49-59.e1. 10.1016/j.jclinepi.2019.03.008

Hsu, C.-C., & Sandford, B. A. (2007). The Delphi technique: Making sense of consensus. Practical Assessment, Research, and Evaluation, 12(1).

Jünger, S., Payne, S. A., Brine, J., Radbruch, L., & Brearley, S. G. (2017). Guidance on Conducting and REporting DElphi Studies (CREDES) in palliative care: Recommendations based on a methodological systematic review. Palliative Medicine, 31(8), 684–706. 10.1177/0269216317690685

Keeney, S., McKenna, H. P., & Hasson, F. (2011). The Delphi technique in nursing and health research. John Wiley & Sons. Kerr, K., Burns, L., Benjamin, S., Joyce, E. M., Singh, J., Ramírez-Bermúdez, J., Stanton, B., & Bell, V. (2025). Unpacking neuropsychiatry and behavioural neurology training: Scoping review of core syllabus components. BJPsych Bulletin, 1–8. 10.1192/bjb.2025.10184

Khachadourian, V., Mahjani, B., Sandin, S., Kolevzon, A., Buxbaum, J. D., Reichenberg, A., & Janecka, M. (2023). Comorbidities in autism spectrum disorder and their etiologies. Translational Psychiatry, 13(1), 71. 10.1038/s41398-023-02374-w

Koh, W. Q., Roes, M., de Vugt, M., Sezgin, D., Gonçalves-Pereira, M., Müller, N., Diaz, A., Casey, D., Ingram, C., Walden, A. K., Thyrian, J. R., Neal, D., Comas, A., & Hopper, L. (2025). What are the unmet needs in people affected by dementia? A scoping review of reviews. Aging & Mental Health, 0(0), 1–20. 10.1080/13607863.2025.2534406

Kosmidis, M. H., Lettner, S., Hokkanen, L., Barbosa, F., Persson, B. A., Baker, G., Kasten, E., Ponchel, A., Mondini, S., Varako, N., Nikolai, T., Jónsdóttir, M. K., Pranckeviciene, A., Hessen, E., & Constantinou, M. (2022). Core Competencies in Clinical Neuropsychology as a Training Model in Europe. Frontiers in Psychology, 13. 10.3389/fpsyg.2022.849151

Krause, C. A., Escamilla, K. J., Hyland, M. T., Gonzalez, A. S., Woods, S. P., Cirino, P. T., Williams, M. W., & Medina, L. D. (2025). An initial review of doctoral clinical neuropsychology coursework syllabi: Common trends and core competencies. Journal of Clinical and Experimental Neuropsychology, 0(0), 1–11. 10.1080/13803395.2025.2494650

Lange, T., Kopkow, C., Lützner, J., Günther, K.-P., Gravius, S., Scharf, H.-P., Stöve, J., Wagner, R., & Schmitt, J. (2020). Comparison of different rating scales for the use in Delphi studies: Different scales lead to different consensus and show different test-retest reliability. BMC Medical Research Methodology, 20(1), 28. 10.1186/s12874-020-0912-8

Murry, J. W. Jr., & Hammons, J. O. (1995). Delphi: A Versatile Methodology for Conducting Qualitative Research. The Review of Higher Education, 18(4), 423–436.

Nelson, A. P., Roper, B. L., Slomine, B. S., Morrison, C., Greher, M. R., Janusz, J., Larson, J. C., Meadows, M.-E., Ready, R. E., Rivera Mindt, M., Whiteside, D. M., Willment, K., & Wodushek, T. R. (2015). Official Position of the American Academy of Clinical Neuropsychology (AACN): Guidelines for Practicum Training in Clinical Neuropsychology. The Clinical Neuropsychologist, 29(7), 879–904. 10.1080/13854046.2015.1117658

Ramírez Bermúdez, J., Castro-Suarez, S., D’Alessio, L., Holguín Lew, J., Makarem Oliveira, L., Sanches Yassuda, M., Santamaría-García, H., Slachevsky, A., Tamayo Agudelo, W., Torales, J., Vera San Juan, N., & Bell, V. (2026). Towards a Latin American neuropsychiatry: Challenges and opportunities. The Lancet Regional Health - Americas, 54, 101322. 10.1016/j.lana.2025.101322

Reiff, C. M., Richman, E. E., Nemeroff, C. B., Carpenter, L. L., Widge, A. S., Rodriguez, C. I., Kalin, N. H., McDonald, W. M., & and the Work Group on Biomarkers and Novel Treatments, a D. of the A. P. A. C. of R. (2020). Psychedelics and Psychedelic-Assisted Psychotherapy. American Journal of Psychiatry. 10.1176/appi.ajp.2019.19010035

Sharma, M., Murphy, R., & Doody, G. A. (2019). Do we need a core curriculum for medical students? A scoping review. BMJ Open, 9(8), e027369. 10.1136/bmjopen-2018-027369

Sheer, D. E., & Lubin, B. (1980). Survey of training programs in clinical neuropsychology. Journal of Clinical Psychology, 36(4), 1035–1040. 10.1002/1097-4679(198010)36:4%253C1035::AID-JCLP2270360439%253E3.0.CO;2-E

Taslim, S., Shadmani, S., Saleem, A. R., Kumar, A., Brahma, F., Blank, N., Bashir, M. A., Ansari, D., Kumari, K., Tanveer, M., Varrassi, G., Kumar, S., & Raj, A. (2024). Neuropsychiatric Disorders: Bridging the Gap Between Neurology and Psychiatry. Cureus, 16(1), e51655. 10.7759/cureus.51655

The Neurological Alliance. (2019). Neuro Patience: Still waiting for improvements in treatment and care: The National Neurology Patient Experience Survey 2018/19: Policy report. https://www.neural.org.uk/wp-content/uploads/2019/07/neuro-patience-2019-1.pdf

Tolchin, B., Hirsch, L. J., & LaFrance, W. C. (2020). Neuropsychiatric Aspects of Epilepsy. Psychiatric Clinics of North America, 43(2), 275–290. 10.1016/j.psc.2020.02.002

Viljoen, C. A., Millar, R. S., Manning, K., & Burch, V. C. (2020). Determining electrocardiography training priorities for medical students using a modified Delphi method. BMC Medical Education, 20(1), 431. 10.1186/s12909-020-02354-4

Wong, D., Pestell, C., Oxenham, V., Stolwyk, R., & Anderson, J. (2024). Competencies unique to clinical neuropsychology: A consensus statement of educators, practitioners, and professional leaders in Australia. The Clinical Neuropsychologist, 38(1), 1–20. 10.1080/13854046.2023.2200035

Woodcock, T., Adeleke, Y., Goeschel, C., Pronovost, P., & Dixon-Woods, M. (2020). A modified Delphi study to identify the features of high quality measurement plans for healthcare improvement projects. BMC Medical Research Methodology, 20(1), 8. 10.1186/s12874-019-0886-6

Zawawi, N. S. M., Aziz, N. A., Fisher, R., Ahmad, K., & Walker, M. F. (2020). The Unmet Needs of Stroke Survivors and Stroke Caregivers: A Systematic Narrative Review. Journal of Stroke and Cerebrovascular Diseases, 29(8). 10.1016/j.jstrokecerebrovasdis.2020.104875

