## Supplementary material for "Developing a Neuropsychiatry Curriculum for Clinical Psychologists and Neuropsychologists: An e-Delphi Study"

**Developing a Neuropsychiatry Curriculum**

**for Clinical Psychologists and Neuropsychologists:**

**An e-Delphi Study**

**Table S1.** Syllabus items rated in e-Delphi round one

| *Syllabus items rated in e-Delphi round one* | *Median* | *MDM* |
| --- | --- | --- |
| Functional neurological disorders (FND) | 3 | 0 |
| Neuropsychiatry of dementia | 3 | 0 |
| Formulating neuropsychological assessments for neuropsychiatric syndromes | 3 | 0 |
| Neuropsychiatry of malnutrition | 2 | 0 |
| Philosophy of neuropsychiatry | 2 | 0 |
| Secondary ('organic') psychiatric syndromes associated with neurological disorders | 3 | 0.05 |
| Neuropsychiatric syndromes of stroke | 3 | 0.05 |
| Metabolic disorders | 2 | 0.05 |
| History of neuropsychiatry | 2 | 0.05 |
| Vestibular disorders | 2 | 0.05 |
| Alcohol-related brain damage and other alcohol-related syndromes | 3 | 0.11 |
| Movement disorders including Parkinson’s disease, Huntington’s disease, and tic disorders | 3 | 0.11 |
| Frontal lobe syndromes | 3 | 0.11 |
| Functional cognitive disorder | 3 | 0.11 |
| Catatonia | 2 | 0.11 |
| Disorders of consciousness including vegetative and minimally conscious states | 2 | 0.11 |
| Understanding the placebo effect | 2 | 0.11 |
| Role of gut-brain axis in neuropsychology | 2 | 0.11 |
| Neuropsychiatry of multiple sclerosis | 3 | 0.16 |
| Challenging behaviour and aggression in people with neuropsychiatric disorders | 3 | 0.16 |
| Post-concussion syndromes | 3 | 0.16 |
| Personality changes in neuropsychiatric disorders | 3 | 0.16 |
| Cultural issues in assessment and practice | 3 | 0.16 |
| Cognitive difficulties in psychiatric disorders | 3 | 0.16 |
| CBT for functional neurological disorder | 3 | 0.16 |
| Apathy and amotivation | 3 | 0.16 |
| Functional neuroimaging | 2 | 0.16 |
| Interpreting structural brain scans | 2 | 0.16 |
| Endocrine disorders | 2 | 0.16 |
| Headache and migraine | 2 | 0.16 |
| Folie à deux and shared delusional disorders | 2 | 0.16 |
| Global neuropsychiatry | 2 | 0.16 |
| Mental capacity and safeguarding law for people with neuropsychiatric disorders | 3 | 0.21 |
| Psychotherapy for neuropsychiatric patients | 3 | 0.21 |
| Effects of common medications used in neuropsychiatry | 3 | 0.21 |
| Delusional misidentification syndromes | 2 | 0.21 |
| Neuropsychiatric syndromes associated with adverse effects of medications | 3 | 0.26 |
| Neuropsychiatric consequences of neurosurgery | 3 | 0.26 |
| Illicit drug-related neuropsychiatric syndromes | 3 | 0.26 |
| Confabulation | 3 | 0.26 |
| Epigenetics of mental health | 2 | 0.26 |
| Neuroinfections including HIV, meningitis, and encephalitis | 3 | 0.32 |
| Neuropsychiatry of brain tumours and neuro-oncology | 3 | 0.32 |
| The function and purpose of the neurological examination | 3 | 0.32 |
| Family interventions for neuropsychiatric disorders | 3 | 0.32 |
| Anosognosia after acquired brain injury | 3 | 0.32 |
| Impulse control disorders | 3 | 0.32 |
| Transient global amnesia | 2 | 0.32 |
| Psychological interventions for tics and Tourette’s syndrome | 2 | 0.32 |
| Genetic neuropsychiatric syndromes | 2 | 0.32 |
| Mass psychogenic illness | 2 | 0.32 |
| Hypnosis | 2 | 0.32 |
| Autoimmune and neuroinflammatory disorders | 3 | 0.37 |
| Positive behaviour support for challenging behaviour | 3 | 0.37 |
| Neuropsychiatric effects of neurotoxicological syndromes | 2 | 0.37 |
| Psychedelic-assisted psychotherapy | 2 | 0.37 |
| Art therapy techniques and applications | 1 | 0.37 |
| Delirium | 3 | 0.42 |
| Focal brain syndromes | 3 | 0.42 |
| Illness exaggeration and simulation in context | 2 | 0.42 |
| Factitious disorders | 2 | 0.42 |
| Sleep-wake disorders | 2 | 0.42 |
| Chronic fatigue syndromes | 2 | 0.42 |
| Hydrocephalus | 2 | 0.42 |
| Evolutionary models of neuropsychiatric disorders | 2 | 0.42 |
| Mitochondrial disorders | 2 | 0.42 |
| Body integrity identity disorder | 1 | 0.42 |
| Interpreting blood test results | 1 | 0.42 |
| Interpreting genetic tests | 1 | 0.47 |
| Motor neuron disease (MND) | 3 | 0.47 |
| Emotionalism, pseudobulbar affect, or emotional expression that doesn't match experienced emotions | 3 | 0.47 |
| Adult ADHD | 3 | 0.47 |
| Medicolegal issues in neuropsychiatry | 2 | 0.47 |
| Cerebellar cognitive affective syndrome | 3 | 0.53 |
| Disorders of smell | 1 | 0.53 |
| Interpreting EEG results | 1 | 0.53 |
| Intellectual and learning disabilities of neurodevelopment | 3 | 0.58 |
| Transient loss of consciousness | 3 | 0.58 |
| Synaesthesia | 1 | 0.58 |

**Table S2.** Syllabus items removed after round one (met rejection threshold criteria)

| Syllabus items removed after round one | Median | MDM | % Scored 1 |
| --- | --- | --- | --- |
| Synaesthesia | 1 | 0.58 | 53% |
| Disorders of smell | 1 | 0.53 | 53% |
| Interpreting EEG results | 1 | 0.53 | 58% |
| Interpreting genetic tests | 1 | 0.47 | 53% |
| Body integrity identity disorder | 1 | 0.42 | 58% |
| Interpreting blood test results | 1 | 0.42 | 68% |
| Art therapy techniques and applications | 1 | 0.37 | 63% |

**Table S3.** Syllabus items suggested by panel members during e-Delphi round one

*Suggested items from free-text box during round one*

Neuropsychiatric disorders of epilepsy

Side effects of commonly prescribed neuropsychiatric medications

Neuropsychiatric assessment and treatment of low mood

Qualitative aspects of performance on neuropsychological tests in neuropsychiatric syndromes

The interplay between mental capacity act and mental health act

Subtle/attenuated presentations of neuropsychiatric phenomena

Psychodynamic psychotherapy

Physiotherapy

Neuropsychiatry of COVID-19

Frontal lobe paradox and mental capacity

Autism

Visual hallucinations

Physical features of mental health problems (e.g., psychomotor agitation, sympathetic nervous system)

Driving in context of seizures and non-epileptic attack disorder

Multi-disciplinary working for neuro-rehabilitation goals

Neuropsychological aspects of medical conditions

Neurological / neuropsychological consequences of common psychiatric conditions

**Table S4.** Syllabus item that met consensus criteria during e-Delphi round one

| *Syllabus items that met consensus threshold in round one* | *Median* | *MDM* |
| --- | --- | --- |
| Neuropsychiatry of traumatic brain injury | 3 | 0 |
| Functional neurological disorders | 3 | 0 |
| Neuropsychiatry of dementia | 3 | 0 |
| Formulating neuropsychological assessments for neuropsychiatric syndromes | 3 | 0 |
| Secondary ('organic') psychiatric syndromes associated with neurological disorders | 3 | 0.05 |
| Neuropsychiatric syndromes of stroke | 3 | 0.05 |
| Alcohol-related brain damage and other alcohol-related syndromes | 3 | 0.11 |
| Movement disorders including Parkinson’s disease, Huntington’s disease, and Tic disorders | 3 | 0.11 |
| Frontal lobe syndromes | 3 | 0.11 |
| Functional cognitive disorder | 3 | 0.11 |
| Neuropsychiatry of multiple sclerosis | 3 | 0.16 |
| Challenging behaviour and aggression in people with neuropsychiatric disorders | 3 | 0.16 |
| Post-concussion syndromes | 3 | 0.16 |
| Personality changes in neuropsychiatric disorders | 3 | 0.16 |
| Cultural issues in assessment and practice | 3 | 0.16 |
| Cognitive difficulties in psychiatric disorders | 3 | 0.16 |
| CBT for functional neurological disorder | 3 | 0.16 |
| Apathy and amotivation | 3 | 0.16 |
| Neuropsychiatry of malnutrition | 2 | 0 |
| Philosophy of neuropsychiatry | 2 | 0 |
| Metabolic disorders | 2 | 0.05 |
| History of neuropsychiatry | 2 | 0.05 |
| Vestibular disorders | 2 | 0.05 |
| Catatonia | 2 | 0.11 |
| Disorders of consciousness including vegetative and minimally conscious states | 2 | 0.11 |
| Understanding the placebo effect | 2 | 0.11 |
| Role of gut-brain axis in neuropsychology | 2 | 0.11 |
| Functional neuroimaging | 2 | 0.16 |
| Interpreting structural brain scans | 2 | 0.16 |
| Endocrine disorders | 2 | 0.16 |
| Headache and migraine | 2 | 0.16 |
| Folie à deux and shared delusional disorders | 2 | 0.16 |
| Global neuropsychiatry | 2 | 0.16 |

**Table S5.** Syllabus items rated in e-Delphi round two

| *Syllabus items rated in e-Delphi round two* | *Median* | *MDM* |
| --- | --- | --- |
| Formulating neuropsychological assessments for neuropsychiatric syndromes | 3 | 0.06 |
| Psychotherapy for neuropsychiatric patients | 3 | 0.12 |
| Qualitative aspects of performance on neuropsychological tests in neuropsychiatric syndromes | 3 | 0.18 |
| Confabulation | 3 | 0.24 |
| Neuropsychiatric disorders of epilepsy | 3 | 0.24 |
| Illicit drug-related neuropsychiatric syndromes | 3 | 0.35 |
| Emotionalism, pseudobulbar affect or emotional expression that doesn't match experienced emotions | 3 | 0.41 |
| Anosognosia after acquired brain injury | 3 | 0.41 |
| Frontal lobe paradox and mental capacity | 3 | 0.41 |
| Neuropsychological aspects of medical conditions | 3 | 0.41 |
| Neurological / neuropsychological consequences of common psychiatric conditions | 3 | 0.41 |
| Impulse control disorders | 3 | 0.47 |
| Focal brain syndromes | 3 | 0.47 |
| Mental capacity and safeguarding law for people with neuropsychiatric disorders | 3 | 0.47 |
| Family interventions for neuropsychiatric disorders | 3 | 0.47 |
| Positive behaviour support for challenging behaviour | 3 | 0.47 |
| Multi-disciplinary working for neuro rehabilitation goals | 3 | 0.47 |
| Neuroinfections including HIV, meningitis, and encephalitis | 3 | 0.53 |
| Delirium | 3 | 0.53 |
| Adult ADHD | 3 | 0.53 |
| Illness exaggeration and simulation in context | 3 | 0.53 |
| Chronic fatigue syndromes | 3 | 0.59 |
| The interplay between mental capacity act and mental health act | 3 | 0.59 |
| Intellectual and learning disabilities of neurodevelopment | 3 | 0.65 |
| Neuropsychiatry of brain tumours and neuro-oncology | 2 | 0.47 |
| Transient global amnesia | 2 | 0.47 |
| Neuropsychiatric assessment and treatment of low mood | 2 | 0.47 |
| Visual hallucinations | 2 | 0.47 |
| Autoimmune and neuroinflammatory disorders | 2 | 0.41 |
| Neuropsychiatric consequences of neurosurgery | 2 | 0.41 |
| Transient loss of consciousness | 2 | 0.41 |
| Effects of common medications used in neuropsychiatry | 2 | 0.41 |
| Motor neuron disease (MND) | 2 | 0.41 |
| Hydrocephalus | 2 | 0.41 |
| Psychological interventions for Tics and Tourette’s syndrome | 2 | 0.41 |
| Side effects of commonly prescribed neuropsychiatric medications | 2 | 0.41 |
| The function and purpose of the neurological examination | 2 | 0.35 |
| Neuropsychiatric syndromes associated with adverse effects of medications | 2 | 0.35 |
| Cerebellar cognitive affective syndrome | 2 | 0.35 |
| Factitious disorders | 2 | 0.35 |
| Delusional misidentification syndromes | 2 | 0.35 |
| Autism | 2 | 0.35 |
| Physical features of mental health problems (e.g., psychomotor agitation, sympathetic nervous system) | 2 | 0.35 |
| Sleep-wake disorders | 2 | 0.29 |
| Neuropsychiatric effects of neurotoxicological syndromes | 2 | 0.29 |
| Genetic neuropsychiatric syndromes | 2 | 0.29 |
| Medicolegal issues in neuropsychiatry | 2 | 0.24 |
| Neuropsychiatry of COVID-19 | 2 | 0.24 |
| Driving in context of seizures and non-epileptic attack disorder | 2 | 0.06 |
| Subtle/attenuated presentations of neuropsychiatric phenomena | 2 | 0.12 |
| Mass psychogenic illness | 2 | 0.18 |
| Mitochondrial disorders | 2 | 0.24 |
| Psychodynamic psychotherapy | 2 | 0.29 |
| Hypnosis | 2 | 0.35 |
| Psychedelic-assisted psychotherapy | 2 | 0.41 |
| Evolutionary models of neuropsychiatric disorders | 2 | 0.41 |
| Physiotherapy | 2 | 0.41 |
| Epigenetics of mental health | 2 | 0.47 |
